# Associations of Healthy Lifestyles with All-Cause Mortality Among Individuals with Osteoarthritis: Results from UK Biobank and US NHANES

**DOI:** 10.1101/2023.04.23.23288990

**Authors:** Tianxiang Fan, Muhui Zeng, Xiaofeng Fang, Shibo Chen, Hao Yang, Yujie Zhang, Ye Li, Peihua Cao, Zhiqiang Wang, Yan Zhang, Qian Yang, Haowei Chen, Weiyu Han, Lijun Lin, Hongbo Guo, David J Hunter, Changhai Ding, Siu Ngor Fu, Zhaohua Zhu

## Abstract

**OBJECTIVE:** To investigate the association of both individual and combined healthy lifestyle factors with the risk of all-cause mortality among patients with osteoarthritis (OA).

**DESIGN:** Prospective population-based cohort study.

**SETTING:** UK biobank and US National Health and Nutrition Examination Survey (US NHANES, 2007-2018)

**PARTICIPANTS:** 104, 142 UK participants with OA aged 39-72 years and 3, 472 US participants with OA aged 20-80 years.

**EXPOSURES:** Individual healthy lifestyle factors and a combined healthy lifestyle score were constructed from body mass index (BMI) and self-reported information on diet, sleep duration, physical activity, sedentary time, social connection, smoking and alcohol drinking.

**MAIN OUTCOME MEASURES:** All-cause mortality was the primary outcome in both studies. Secondary outcomes included cause-specific mortalities (cardiovascular, cancer, digestive and respiratory). Hazard ratios were adjusted for age, sex, economic situation, race, education and employment (UK biobank only).

**RESULTS:** UK Biobank documented 9,914 deaths during a median follow-up of 12.7 years, and US NHANES documented 463 deaths during a mean follow-up of 6.01 years. For all-cause mortality using restricted cubic spline graph (RCS) models, sleep duration had a U-shaped (with a nadir at 7 hours/day), moderate physical activity (MPA) had an L-shaped (with a turning point at 550 minutes/week), while BMI, vigorous physical activity (VPA) and sedentary time had J-shaped (with turning points at 28 kg/m^2^, 240 minutes/week and 5 hours/day, respectively) associations in the UK biobank. Similar results were observed in US NHANES. In multivariable Cox models, each healthy lifestyle factor was significantly associated with all-cause mortality (hazard ratio [HRs] range 0.49 to 0.84 for UK biobank, and 0.26 to 0.73 for US NHANES), and HRs (95% CI) for associations with combined healthy lifestyle score (scoring 6-8 vs. 0-2) were 0.38 (0.35, 0.41) in UK biobank and healthy lifestyle score (scoring 5-7 vs. 0-1) were 0.20 (0.13, 0.31) in US NHANES for all-cause mortality. The results for cause-specific mortality were largely similar and consistent across two cohorts.

**CONCLUSIONS:** The nonlinear relationships suggested patients with OA had the lowest risk of all-cause mortality when BMI was 28 kg/m^2^, sleep was 7 hours/day, VPA was 240 minutes/week, sedentary time was less than 5 hours/day, MPA was more than 550 minutes/week. The newly constructed healthy lifestyle score for OA population was associated with a significantly lower risk of all-cause mortality.

**WHAT IS ALREADY KNOWN ON THIS TOPIC:** Healthy lifestyles are thought to reduce the risk of multiple causes of mortality in the general population.

People with osteoarthritis (OA) are at higher risk of mortality than the general population. However, evidence of associations between combined healthy lifestyle and risks of all-cause and cause-specific mortality among OA patients are lacking.

Whether and what kind of healthy lifestyles in patients with OA could offset the risk of mortality are unknown.

**WHAT THIS STUDY ADDS:** By using two nationwide cohort studies in UK and US, a comprehensive healthy lifestyle pattern that integrates 8 or 7 lifestyle factors was established among OA individuals for the first time.

People who had the highest combined healthy lifestyle score were significantly associated with 62% to 80% lower risk of all-cause mortality compared with those who had the lowest score in both the UK and US OA populations.

## Introduction

Osteoarthritis (OA) is the commonest form of arthritis, affecting approximately 7% of the global population, with particularly high prevalence in those of advanced age.[1] Globally, the prevalence of OA increased by 113.25% from 247.51 million in 1990 to 527.81 million in 2019, in part due to the influence of accelerated aging and obese population.[2] The mortality risk was significantly higher among people with OA than among the general population.[3] A large population-based cohort study found that knee or hip OA participants had a 1.55 standardized mortality ratio of all-cause and 1.38 to 2.28 of cause-specific mortalities than the general population over a median of 14-years follow-up.[4] While OA may not be a direct cause of death, factors such as comorbidities, physical inactivity and obesity may contribute to the increased risk of all-cause mortality in the OA population.[4–6] Thus, it is crucial to explore strategies to increase life expectancy in individuals with OA.

It is widely acknowledged that healthy lifestyles can impact health and lower mortality risk. [7–12] Some previous studies have examined the effects of individual or several lifestyle factors on mortality in the OA population.[5,13–16] However, significant gaps remain. First, while previous studies have identified lifestyles such as physical inactivity, obesity, and unrefreshed sleep as contributing to increased mortality among OA patients, the optimal degree of lifestyles for reducing mortality in the OA population is still unclear. Second, previous research only explored a limited range of lifestyle factors and disregarded emerging factors such as sedentary behavior, social connection and common risk factors such as alcohol drinking and smoking when evaluating lifestyle scores for mortality risk in individuals with OA.[9] Third, How much an overall healthy lifestyle can reduce the risk of mortality in OA patients remains unknown.

Therefore, this study aimed to investigate the associations of both individual and combined healthy lifestyle factors using body mass index (BMI) and self-reported information on diet, sleep duration, physical activity, sedentary time, social connection, smoking and alcohol drinking with the risk of all-cause and cause-specific mortality among participants with OA using data from the UK biobank and the US National Health and Nutrition Examination Survey (US NHANE, 2007-2018).

## Methods

### Study Populations

We used data from the UK Biobank (application number 67654) and US NHANES (2007-2018). UK Biobank included 502,411 middle-aged (37-73 years) adults from 22 sites across England, Wales, and Scotland with baseline measures collected between 2006 and 2010 and with data linked to mortality records.[17] UK Biobank was approved by the National Health Service (NHS) National Research Ethics Service (16/NW/0274, ethics approval for UK Biobank studies)[18]. After excluding 1298 participants who withdrew from the study and 286 participants who died in the first 2-year follow up, we included 104,142 participants with OA in this study. The US NHANES is a nationally representative study to assess health and nutritional status of the noninstitutionalized civilian population in the United States.[19] We utilized data from six cycles of the NHANES, which was conducted from 2007 to 2018. The US NHANES sample included 34,692 adults aged 20 years or older. After excluding 114 participants who withdrew from the survey and 161 who died in the first 2-year follow-up, a total of 3472 participants diagnosed with OA were included in the analysis (Figure 1).

**Figure 1.**
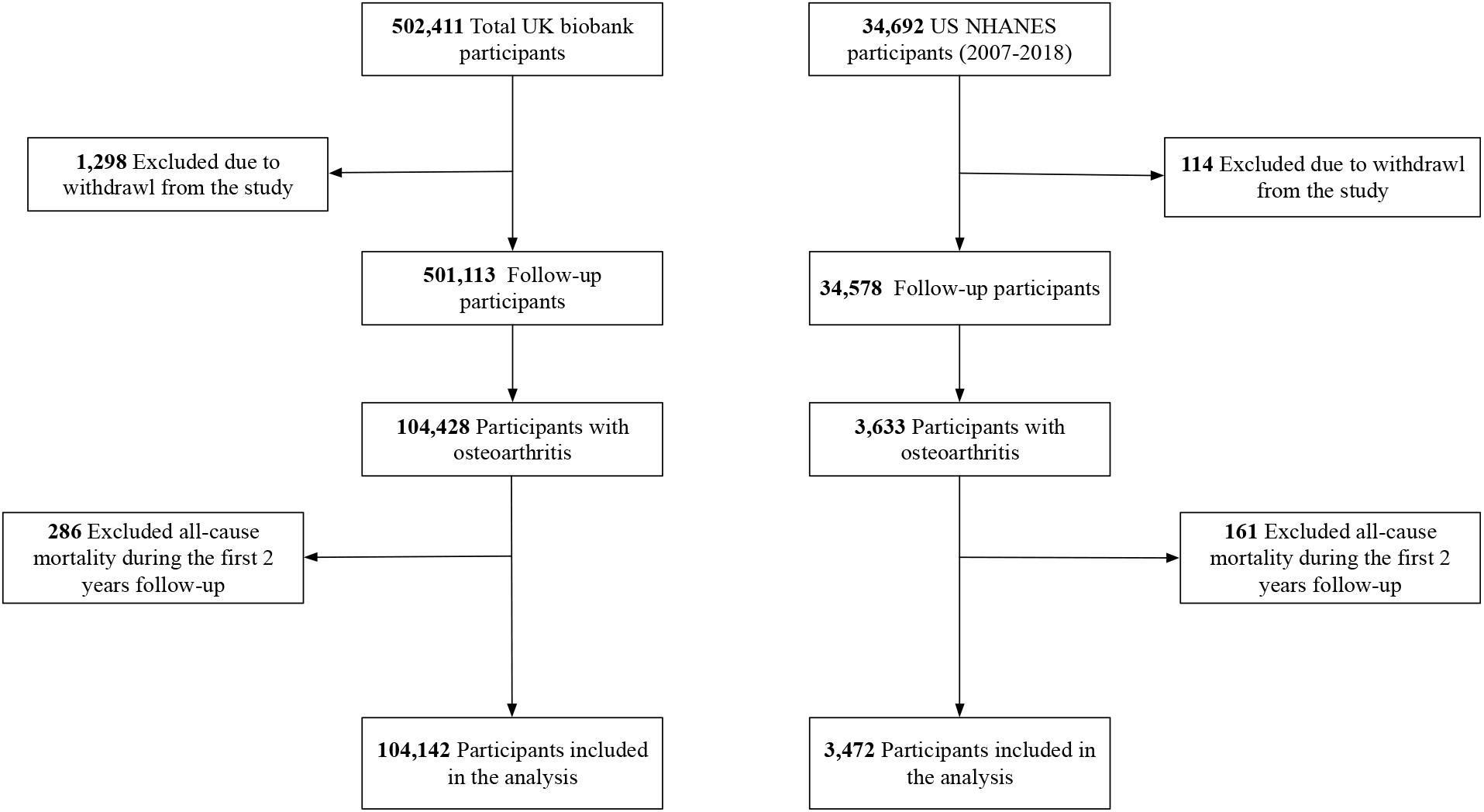
Flow chart of participant enrolment.

### Assessment of Lifestyle factors

We considered eight modifiable lifestyle factors that had previously been interrelated and associated with mortality, including physical activity (PA), BMI, sedentary behavior, social connection, sleep duration, smoking, diet, alcohol consumption and to generate a healthy lifestyle score, because they had previously been reported to be associated with mortality[9,20]. Supplementary table 1 shows deifinitions of each healthy lifesty. In the UK biobank, PA was assessed using the questionnaires of frequency and duration of moderate physical activity (MPA, Field 884 and 894) and vigorous physical activity (VPA, Field 904 and 914) on a typical day/week over the past four weeks. In the US NHANES, participants were queried about their participation in both moderate-(PAQ665 and PAQ670) and vigorous-(PAQ655 and PAD660) intensity recreational and work activities (PAQ625, PAD630, PAQ610 and PAD615) during a typical week. The duration of MPA was calculated as the sum of the minutes spent engaging in moderate-intensity recreational activities and moderate-intensity work activities.[21] Similarly, the minutes of VPA was determined by adding together the time spent participating in vigorous-intensity recreational activities and vigorous-intensity work activities.[21]

According to a modified version of the International Physical Activity Questionnaire (IPAQ) [22,23], duration of PA less than 10 minutes per day for any category was recoded to 0 minutes. Participants with a duration of any type of PA greater than 960 minutes per day were excluded from the analysis as unreasonably high data were outliers. According to the IPAQ guidelines, moderate and vigorous time variables exceeding 240 minutes per day were truncated to equal 240 minutes per day in a new variable. These data processing rules will ensure that highly active people remain highly active, while decreasing the chances that less active individuals are coded as highly active.[24] The total PA score was assigned one point for each condition met (MPA ≥ 550 minutes/week, VPA between 100 minutes/week and 500 minutes/week), which ranged from 0 to 2. Total PA was reclassified in binary as low PA group (0 points) and healthy PA group (1-2 points).

BMI (Field 21001 in UK biobank, BMXBMI in NHANES) value is constructed from height and weight measured during the initial Assessment Centre visit. BMI between 26-30 kg/m^2^ was classified as low-risk BMI.

We used the sum time of time spent watching television (Field 1070), time spent using the computer (Field 1080, not including using a computer at work) and time spent driving (Field 1090) as the proxy for total sedentary time in the UK biobank. In the US NHANES, the sedentary time (PAD680) was self-reported by participants answering “On a typical day, how much time do you usually spend sitting at school, at home, getting to and from places, or with friends, including time spent sitting at a desk, traveling in a car or bus, reading, playing cards, watching television, or using a computer?”. Total sedentary time of more than 24 hours per day was removed, while sedentary time values over 16 hours were winsorized.[25] Sedentary scores were defined as less sedentary time (less than 3 hours/day), moderately sedentary time (3-5 hours/day) and long sedentary time (more than 5 hours/day).[26]

In the UK biobank, information on the number in the household, frequency of friend/family visits, and participation in leisure/social activity was used to evaluate the social connection level.[27] Social connection score was summed to calculate an overall score ranging from 0 to 3, defining as most isolated if they scored 0 or 1 (since few individuals had scores of 0), moderately isolated if they scored 2, and least isolated if they scored 3. Achievement of at least 2 of 3 social connection components was considered as a healthy social connection, and less than 2 was considered a poor social connection. We did not construct a social connection score in US NHANES because of no such data.

In the UK biobank, participants were asked the question, “About how many hours of sleep do you get in every 24 hours?” to quantify sleep duration (Field 1160). In US NHANES, participants were asked the question, “How much sleep do you usually get at night on weekdays or workdays?” to quantify sleep duration (SLD010H and SLD012). According to RCS, the total sleep hours per day were categorized into three groups: low (less than 7 hours/day), moderate (7-8 hours/day), and high (more than 8 hours/day). Sleep duration between 7 hours/day to 8 hours/day is regarded as a low-risk level.

In the UK biobank, a healthy diet was defined as an adequate intake of at least 3 of 5 food groups recommended (Supplementary table 2) as dietary priorities for cardiometabolic health: increased consumption of fruits, whole grains, fish and reduced consumption of refined grains, processed meats.[28] Dietary intakes used in the US NHANES study were obtained using data from the two 24-hour dietary recalls. We used the latest iteration of the Healthy Eating Index (HEI) 2015.[29] It consists of 13 components that are scored based on energy-adjusted food and nutrient intakes. Of the 13 components, 9 assess dietary adequacy (total fruits, whole fruits, total vegetables, greens and beans, whole grains, dairy, total protein foods, and fatty acids) and 4 assess moderation (refined grains, sodium, added sugar and saturated fats). According to quantile, the HEI was categorized from 0-5 and at least 3 of 5 was considered as a healthy diet.

No current smoking (Field 20116 in the UK) was classified as low risk. In US NHANES, no current smoking (SMQ020 and SMQ040) was defined as never smoking (smoked less than 100 cigarettes in life) or former smoking (smoked more than 100 cigarettes in life and smoke not at all now). Low-risk alcohol consumption was defined as moderate drinking (no more than two drinks/day; one drink is measured as 8 g ethanol in the U.K, 12 g ethanol in the US) on a relatively regular frequency. Participants who reported no drinking or drinking only on special occasions were regarded as nonregular drinkers.

Each lifestyle factor was coded as 1 (protective) or 0 (referent), and the sum of these scores together gave a newly constructed score of 0 to 8 in the UK biobank and 0 to 7 in the US NHANES (US NHANES lacks information of social connection), with higher scores indicating a healthier lifestyle.

### Disease diagnosis

In the UK biobank, International Classification of Diseases, tenth Revision (ICD-10) codes were used. OA was identified through hospital inpatient records that were classified based on M15, M16, M17, M18, M19, M471, M472, M478, M479, and M480. Cancer diagnosis codes were C00-C97; Cardiovascular Disease (CVD) diagnosis codes were I20-I25, I60-I64 I10-I13; Respiratory disease diagnosis codes were J40-J44, J47; Digestive disease diagnosis codes were K70-K77. In US NHANES, OA was identified by an answer of “yes” to the question “Has a doctor or other health professional ever told you that you had arthritis?”(MCQ195, MCQ191 and MCQ190) and “Osteoarthritis” to the question “Which type of arthritis was it?”(MCQ160A). Cancer diagnoses were defined as self-reported questionnaires “Have you ever been told by a doctor or other health professional that you had cancer or a malignancy of any kind?”(MCQ220). Chronic obstructive pulmonary disease was defined based on Post-Bronchodilator FEV1 / FVC < 0.7 or self-reported emphysema diagnosis “Has a doctor or other health professional ever told you that you had emphysema?” (MCQ160G) or use drug: selective phosphodiesterase-4 inhibitors, mast cell stabilizers, leukotriene modifiers, inhaled corticosteroids.

### Outcome ascertainment

In the UK biobank, date and cause of death information were obtained from death certificates held within the National Health Service Information Centre (England and Wales) and the National Health Service Central Register Scotland (Scotland). Person-time was calculated from baseline to the occurrence of death or the end of follow-up (England & Wales on 30 September 2021, Scotland on 31 October 2021), whichever came first. The outcomes of this study were all-cause and cause-specific mortality (cancer, CVD, respiratory disease, digestive disease) based on the ICD-10 code (Supplementary table 3);[9] In US NHANES, the NCHS (National Center for Health Statistics) provided mortality data that were linked to the National Death Index through 31 December 2019.[30] The ICD-10 was used to record the underlying cause of death.[19,20] The duration of follow-up was defined as the interval in months from the interview date to the date of death or through December 31, 2019, for those participants who did not experience an event.

### Assessment of Covariates

In the UK biobank, relevant covariates included age at baseline (Field 21022); sex (Field 31); Townsend deprivation index (Field 189, referring to an area-based measure of socioeconomic deprivation) was used for the economic situation; Ethnic background (Field 21000); education (Field 6138), categorized as college or university degree/others; Since UK Biobank did not collect information on the specific occupation of participants at the baseline, we re-categorized employment status (Field 6142) into two groups: employed (including those who were self-employed or in paid employment, as well as retirees, unpaid or voluntary workers, and full or part-time students) and unemployed.

In US NHANES, self-reported covariates included sex (RIDAGEYR), race and ethnicity (non-Hispanic Black, Hispanic, non-Hispanic White, other [American Indian/Native Alaskan/Pacific Islander, Asian, multiracial]) (RIDRETH1), educational attainment (<high school, high school, some college, college graduate or above) (DMDEDUC2), and family poverty income ratio (PIR) (total family income divided by the poverty threshold; <1, 1 ≤ &<3, ≥3) (INDFMPIR) was used for the economic situation.

### Statistical Analyses

Restricted cubic spline (RCS) models fitted for Cox proportional hazards models with three knots to estimate the non-linear associations between BMI, MPA, VPA, sleep duration, sedentary time and all-cause mortality after adjustment for age, sex, economic status, ethnic background, education, employment (only in UK biobank); Potential non-linearity was tested using a likelihood ratio test as described previously.[31] Multivariable Cox proportional hazards models were performed to examine the associations of each lifestyle factor and the combined lifestyle score with all-cause and cause-specific mortality after adjustment for covariates. The results were reported as hazard ratios (HRs) and corresponding 95% confidence intervals (CIs). Missing data were deleted in the Cox analyses. We also conduct stratified analyses by sex (men and women), self-reported race (white and non-white participants), and age groups (<60, and ≥ 60 defined as elders by the World Health Organization[32]) to test the robustness and potential variations in different subgroups.

Several sensitivity analyses have been conducted. First, to address the varying strengths of the relationships between different lifestyle factors and outcomes, a weighed healthy lifestyle score was created.[33] Second, to assess the impact of missing variables, we employed multiple imputations to impute all missing independent variables.[34] Third, restricted the analysis to those aged 40 years or older in US NHANES to coincide with the age distribution in UK Biobank, and the risk of mortality due to lifestyles is relatively lower in younger adults. Since there were only 2 participants under the age of 40 in the UK Biobank, we did not conduct this sensitivity analysis in the UK biobank. Fourth, we excluded individuals with CVD and cancer at baseline, because they may greatly affect lifestyles and mortality.

A two-sided P < 0.05 was considered statistically significant. R software version 4.1.3 was used for statistical analyses and data visualization.

### Patient and public involvement

The analyses were based on existing data from US NHANES and UK Biobank. No patients or participants were involved in the design, recruitment, or conduct of the studies. No patients were asked to advise on interpretation or writing up of results.

## Results

### Population Characteristics

The baseline characteristics of the study population are shown in Table 1. Of the 104,142 participants with OA at baseline (mean age 59.8 years, 42% men), the proportion of unfavorable lifestyles (score from 0 to 3), intermediate lifestyles (score from 4 to 5) and favorable lifestyles (score from 6 to 8) was 31%, 52% and 17% respectively. Among 3,472 participants from US NHANES (mean age 62.51 years, 35% men), 36% had unfavorable lifestyles, 49% had intermediate lifestyles, and 15% had favorable lifestyles. In both cohorts, participants with a lower healthy lifestyle score were more likely to be non-white, less educated, and have lower incomes. CVD, digestive, and respiratory diseases were more prevalent among those with lower healthy lifestyle scores.

**Table 1.**
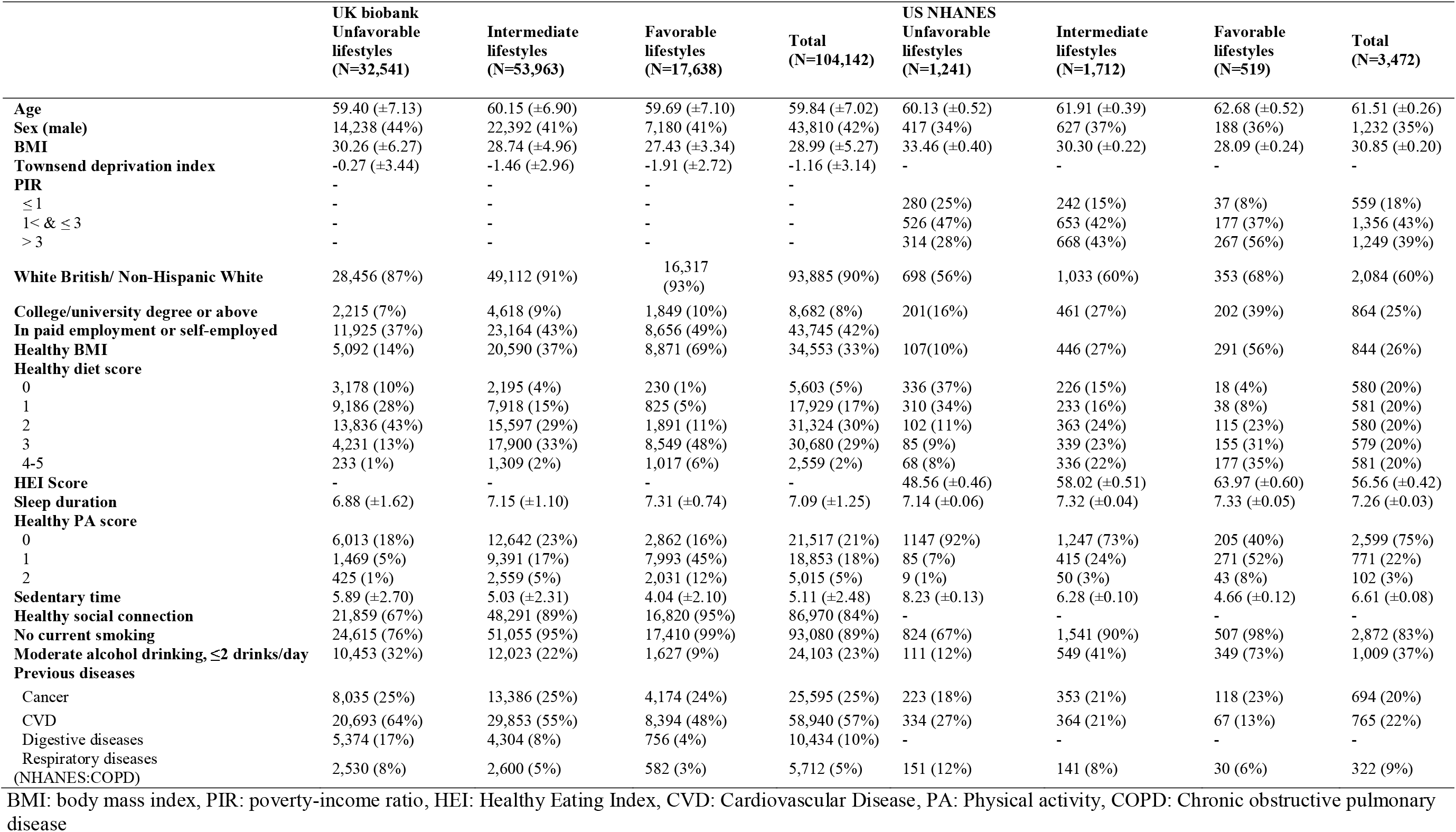
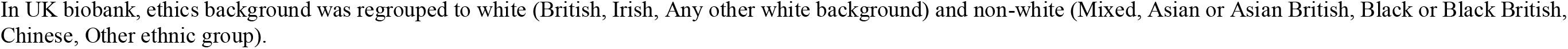
Baseline characteristics of participants from UK Biobank and US NHANES according to healthy lifestyle score.

### Association of Individual and Combined Lifestyle Factors With All-Cause Mortality

UK Biobank documented 9,914 deaths during a median follow-up of 12.7 years, US NHANES documented 463 deaths during a mean follow-up of 6.01 years. Cox regression with RCS was used to explore the relationships between continuous variables (BMI, MPA, VPA, sleep duration and sedentary behavior) and all-cause mortality after adjustment for age, sex, economic conditions, ethnic background, education and employment (only UK biobank). (Figure 2 & Figure 3). The results showed that MPA had a non-linear L-shaped (with a turning point at 550 minutes/week), VPA had a non-linear J-shaped (with turning points 240 minutes/week), association with all-cause mortality in the UK biobank. The healthy MPA were thereby defined as more than 550 minutes/week, because the risk of all-cause mortality was relatively flat after this threshold. The VPA between the two intersection points (100 and 500 minutes/week) of the curve and the reference line was defined as healthy VPA. When compared with the low PA group (with either healthy VPA or healthy MPA), HRs for all-cause mortality were lower for high PA (with healthy vigorous PA and healthy MPA) (HR [95% CI]) = 0.80 [0.71–0.90]) in the UK biobank, and (HR [95% CI]) = 0.26 [0.09–0.81]) in the US NHANES. Given the non-linear J-shaped association between BMI and all-cause mortality in the UK biobank, BMI between 26-30 kg/m^2^ was classified as low-risk BMI. When compared with the high-risk BMI (less than 26 kg/m^2^ or more than 30 kg/m^2^), HRs for all-cause mortality were lower for low-risk BMI (HR [95% CI]) = 0.84 [0.81–0.88]) in the UK biobank, and (HR [95% CI]) = 0.73 [0.57–0.93]) in the US NHANES. According to the non-linear J-shaped association between total sedentary time and all-cause mortality in the UK biobank, total sedentary time less than 5 hours/day was defined as a low-risk level (low sedentary and moderate sedentary). When compared with the long sedentary group, HRs for all-cause mortality were lower for the short sedentary group (HR [95% CI]) = 0.71 [0.67–0.76]) in the UK biobank, and (HR [95% CI]) = 0.49 [0.38–0.63]) in the US NHANES. The non-linear U-shaped association between sleep duration and all-cause mortality in the UK biobank defined sleep duration between 7hours/day to 8hours/day as a low-risk level. When compared with the high-risk sleep (less than 7hours/day or more than 8hours/day), HRs for all-cause mortality were lower for low-risk sleep (HR [95% CI]) = 0.82 [0.79–0.85]) in the UK biobank, and (HR [95% CI]) = 0.76 [0.61–0.95]) in the US NHANES.

**Figure 2.**
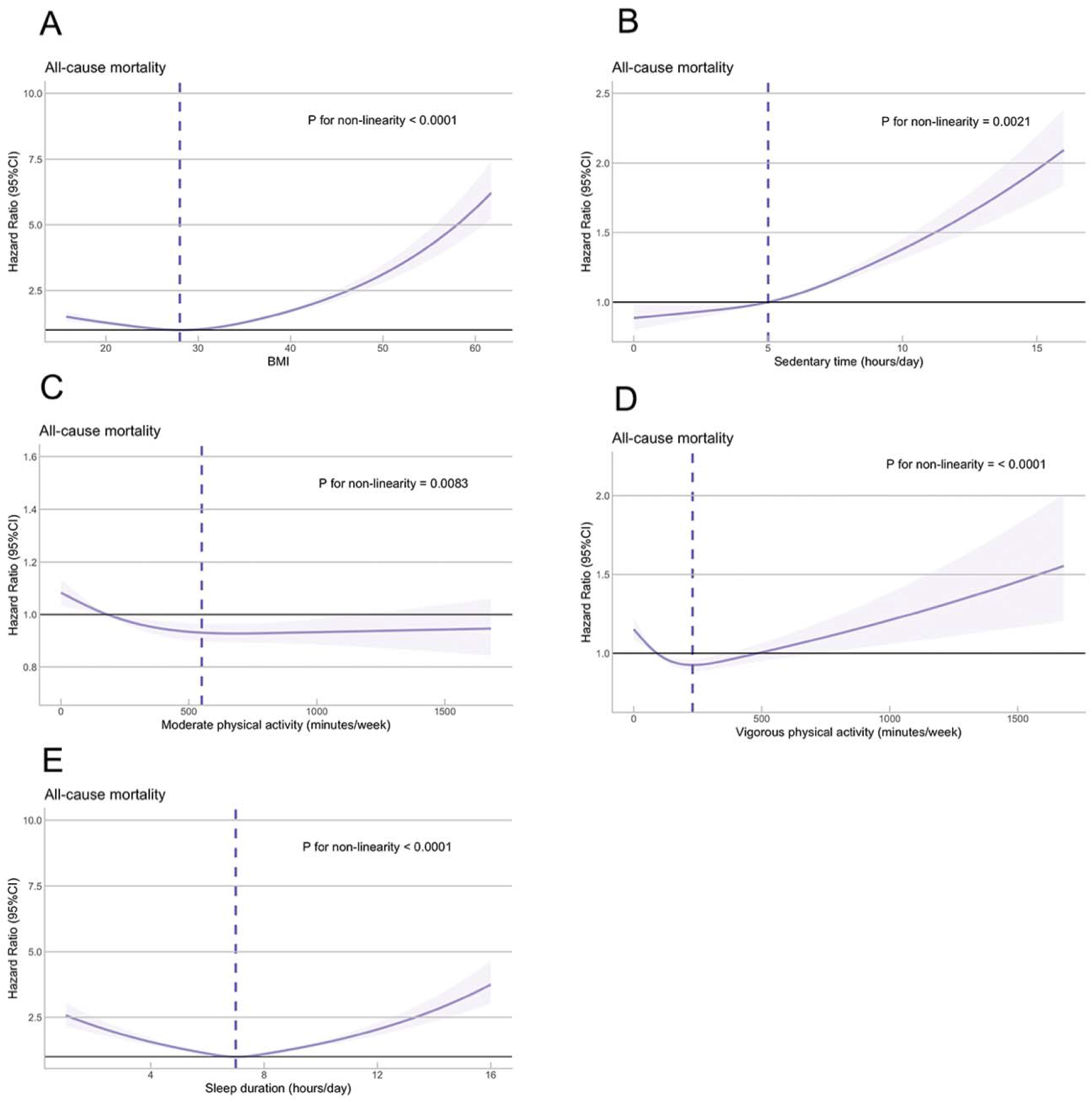
The associations between exposures and all-cause mortality using a restricted cubic spline regression model in UK biobank. Restricted cubic spline models fitted for Cox proportional hazards models with three knots were used. Adjustment for age, sex, Townsend deprivation index, ethnic background, education and employment; The nadirs or turning Points of A, B, C, D and E were 28 kg/m^2^, 550 min/week, 240 min/week, 5 hours/day and 7 hours/day, respectively.

**Figure 3.**
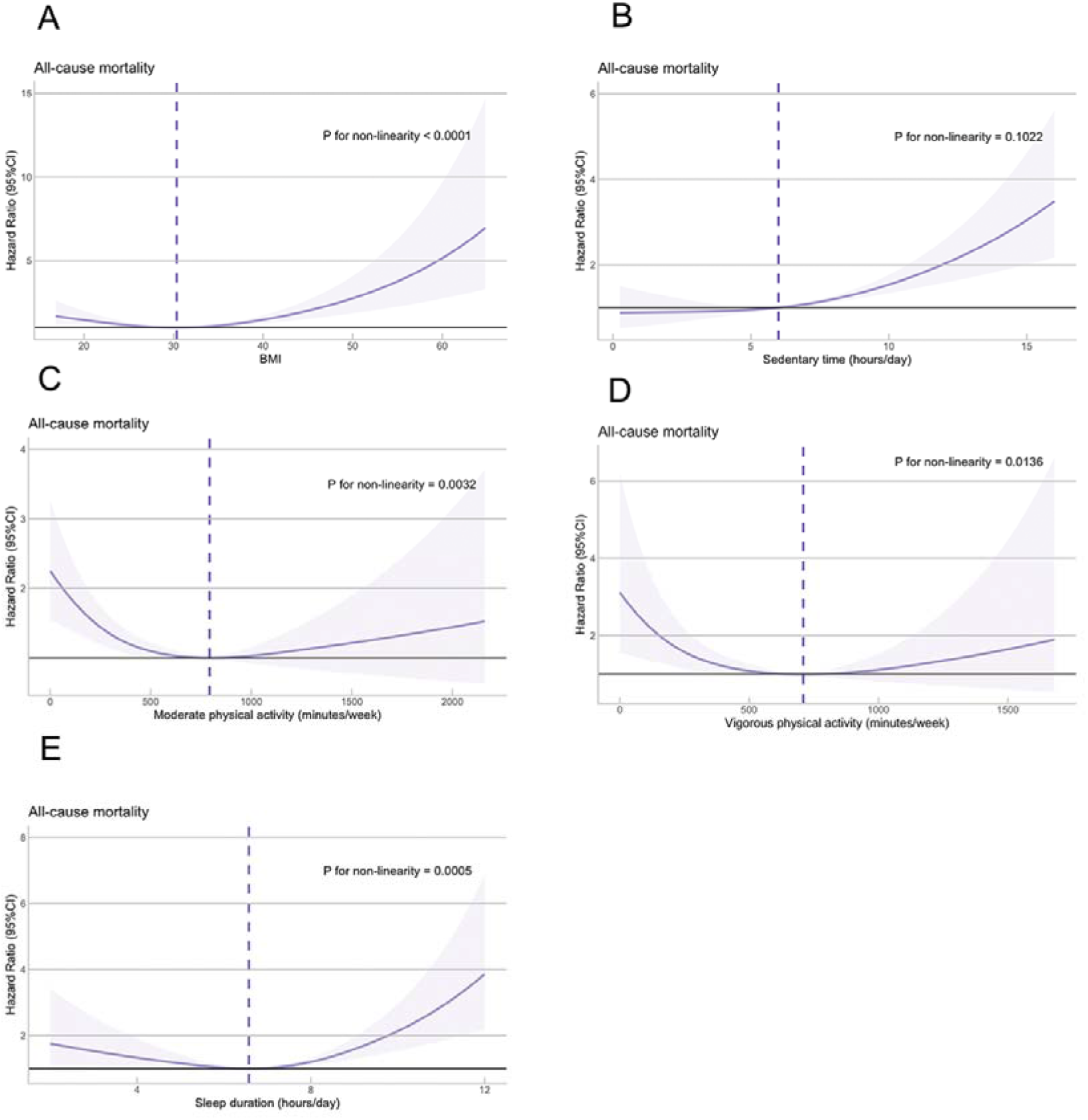
The associations between exposures and all-cause mortality using a restricted cubic spline regression model in US NHANES. Restricted cubic spline models fitted for Cox proportional hazards models with three knots were used. Adjustment for age, sex, PIR, race, and education; The nadirs or turning Points of A, B, C, D and E were 30 kg/m^2^, 792 min/week, 709 min/week, 6 hours/day and 6.6 hours/day, respectively.

We also estimated the all-cause mortality related to non-continuous variables separately. When compared with the healthy diet score 1 group, the risk of all-cause mortality was significantly decreased in the highest diet score group (HR [95% CI]) = 0.66 [0.61–0.73]) in the UK biobank, and (HR [95% CI]) = 0.56 [0.35–0.91]) in the US NHANES. Compared with the current smoking group, no current smoking group had a lower risk of all-cause mortality (HR [95% CI]) = 0.49 [0.47–0.52]) in the UK biobank, (HR [95% CI]) = 0.48 [0.34–0.69]) in the US NHANES. There was a significantly decreased risk of all-cause mortality in the moderate alcohol consumption group compared with non-moderate alcohol consumption group in the UK biobank (HR [95% CI]) = 0.78 [0.75–0.82]), but a non-significant decrease in US NHANES (HR [95% CI]) = 0.86 [0.65–1.14]). The healthy social connection was also found to be significantly associated with reduced risk of all-cause mortality in the UK biobank (HR [95% CI]) = 0.69 [0.66–0.73]). There is no information on social connection in US NHANES. The results of the associations between individual lifestyle and all-cause mortality after further adjustment for other lifestyle factors were shown in Supplementary Table 4.

### Association of Combined Lifestyle Score With All-Cause Mortality

After adjustment for age, sex, economic conditions, ethnic background, education and employment (only UK biobank), each lifestyle factor and combined lifestyle score were significantly associated with all-cause mortality, except moderate drinking in US NHANES (Table 2). Compared with healthy lifestyle scoring 0–2, all-cause mortality risk (HR) for participants having 3, 4, 5, 6-8 healthy lifestyle factors were 0.70 (0.66, 0.74), 0.54 (0.51, 0.57), 0.46 (0.43, 0.50), 0.38 (0.35, 0.41), respectively, in UK biobank. In US NHANES, compared with healthy lifestyle scoring 0–1, all-cause mortality risk for participants having 2, 3, 4, 5-7 healthy lifestyle factors were 0.48 (0.33, 0.69), 0.40 (0.28, 0.58), 0.34 (0.23, 0.50), 0.20 (0.13, 0.31), respectively. Each one-point increase in healthy lifestyle score was significantly associated with lower risk of all-cause mortality [HR = 0.83 (0.82, 0.84), P for trend = 1.49×10^-181^)] in UK biobank and [HR = 0.71 (0.64, 0.78), P for trend = 1.51×10-12)] in US NHANES.

**Table 2.**
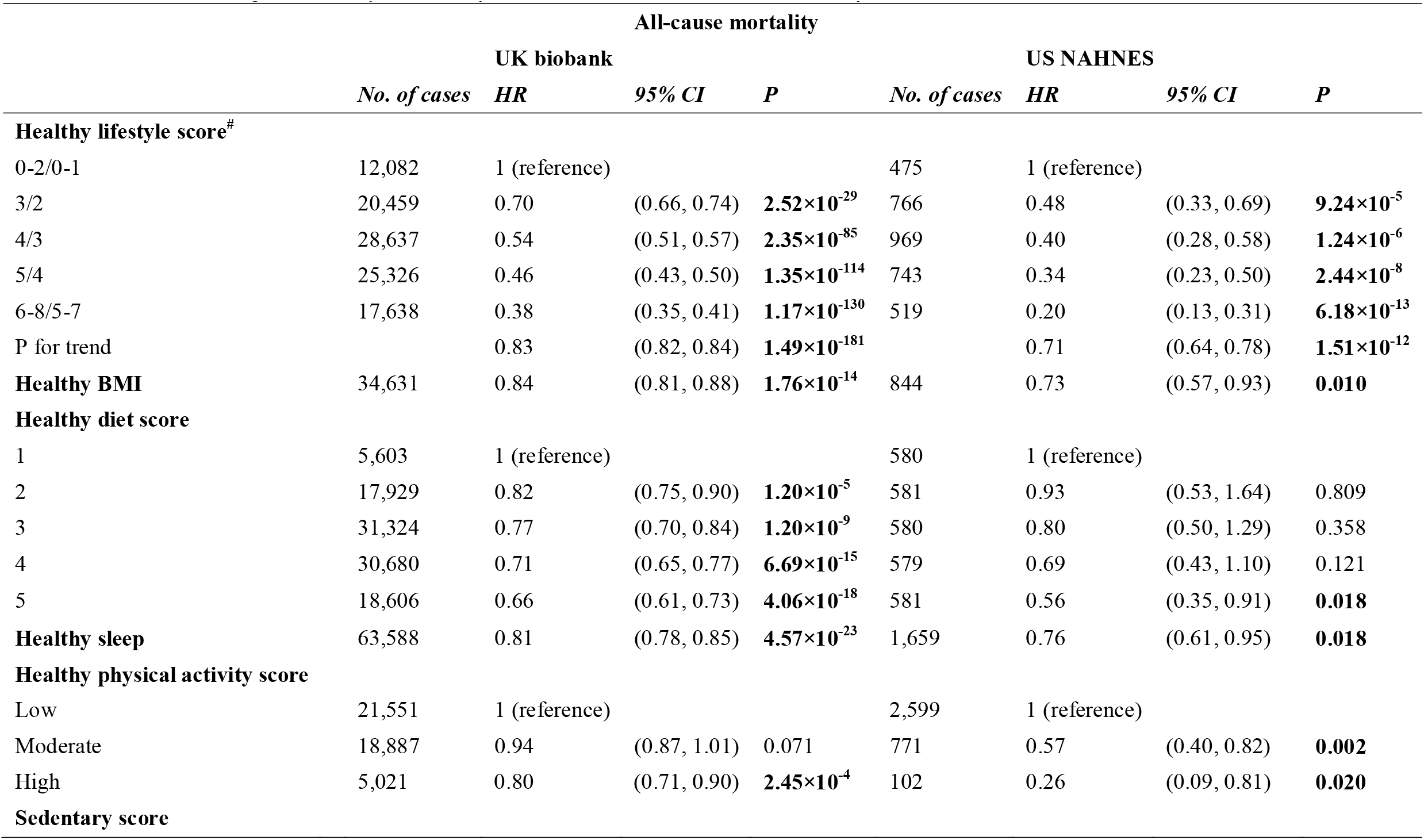
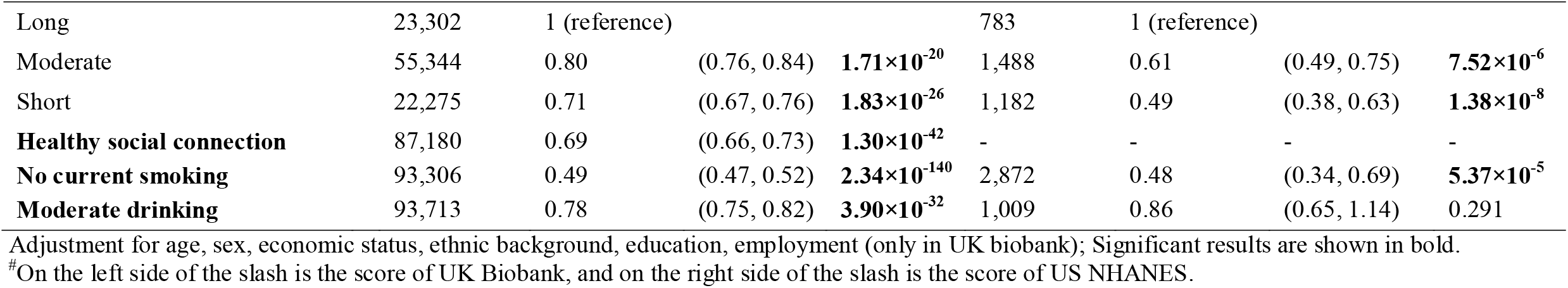
Multivariable cox regression analysis of lifestyle factors in relation to all-cause mortality.

### Sensitivity analysis

In sensitivity analyses, the results were not significantly changed when missing values of covariates and exposure were estimated using multiple imputations (Supplementary Table 5), when the weighted healthy lifestyle score was constructed (Supplementary Table 6), when excluded participants below 40 years old (Supplementary Table 7), and when excluded individuals with CVD or cancer at baseline (Supplementary Table 8 for UK biobank & Supplementary Table 9 for US NHANES).

### Subgroup analysis

Supplementary tables 10, 11, and 12 present results stratified by sex, self-reported race, and age group. The study found that the associations between healthy lifestyle score and all-cause mortality were stronger in females compared to males, stronger in younger adults than older adults and stronger in White individuals compared to non-White individuals in both cohorts (P for interaction <0.05). The healthy lifestyle score was found to decrease the risk of all-cause mortality, and these results were similar to those observed in Table 2.

## Discussion

This study found nonlinear associations between some lifestyle factors and all-cause mortality and defined the optimal range of healthy lifestyles in participants with OA. The significant association between newly defined OA-specific healthy lifestyle scores on reducing the risk of all-cause mortality and cause-specific mortality was confirmed by two large nationwide prospective cohorts in UK and US. Compared to those with zero to two healthy lifestyle factors, those with six to eight healthy lifestyle factors had 62% to 80% lower risk of all-cause mortality in both the UK and US OA populations.

### Comparison with other studies

Evidence suggests that exercise patterns in patients with OA differ significantly from non-OA patients. Most knee OA patients spent hours sedentary (61%), with 4.6 bouts of extended duration (>_30 minutes), and 75% of participants did not perform prolonged PA.[35] A mediation analysis revealed that low physical function might explain 42.9% and 25.0% of the increased risk of all-cause and CVD mortality among women with hip OA, respectively.[15] Our results indicated that engaging in suitable PA can lower the risk of all-cause mortality, which aligns partially with this finding, supporting the importance of PA in OA patients for mortality. Although the latest recommendations from EULAR, ACR and OARSI consider PA as the core treatment for people with OA[36], it is not clear what kind of exercise and how much of it helps to reduce mortality in people with OA. The nonlinear relationships in our study suggested that participants with OA could reduce all-cause mortality risk when they had more than 550 minutes/week of MPA, VPA between 100 to 500 minutes/week and sedentary time less than 5 hours/day. According to the latest PA recommendations for adults in the US and European society, adults should take 150-300 minutes per week of MPA, 75-150 minutes per week of VPA, or a combination of both types of activity.[37–39] Nevertheless, our findings suggest that to minimize the risk of all-cause mortality, individuals with OA should undertake a much greater amount of MPA and VPA than what is recommended for the general public. Notably, our study also suggested VPA of more than 500 minutes/week may even be counterproductive.

High BMI is not only a risk factor for OA disease progression but also for mortality.[40–42] In the general population, relative to normal weight, both grades 2 and 3 obesity were associated with a significantly higher risk of all-cause mortality,[42] and mortality risk was lowest at about 22·5–25 kg/m².[43] Our study suggested that OA patients with BMI between 26-30 had the lowest risk of all-cause mortality compared with others. Healthy dietary component intake for cardiovascular health generally cover ten categories of dietary components.[44,45] While in our study, only 5 of them were found to be significantly associated with all-cause mortality in patients with OA, including fruit, whole grains, shell or fish, refined grains and processed meats.

The optimal sleep duration for health is generally considered 6 to 8 hours, with individuals who sleep for 7 hours having the lowest risk of all-cause mortality.[46,47] In the current study, the nonlinear relationships between sleep duration and all-cause mortality are consistent with previous ones, indicating that both longer and shorter sleep durations are associated with an increased risk of all-cause mortality. Although smoking is a well-established risk factor for various diseases and increased mortality, it has been reported to have a significant positive impact on delaying the progression of OA, which may be due to the effect of nicotine on increasing collagen synthesis in chondrocytes.[48–50] However, our study revealed that OA individuals who currently do not smoke had a lower risk of all-cause mortality, suggesting individuals with OA should quit smoking.

Many previous studies have established a link between diet, alcohol consumption, and health. It has been reported that moderate alcohol consumption and a healthy diet can significantly decrease the risk of all-cause mortality in the general population or in people with diabetes,[9,20,51] and we observed a similar pattern among the OA population. In recent years, social connection has gradually attracted attention as an emerging lifestyle factor, and several studies indicated its relation with all-cause mortality.[52,53] Our research supports previous studies by demonstrating that the mortality rate of OA patients lacking social connection was significantly increased. Individuals suffering from OA frequently exhibit health risk factors that could potentially increase their probability of experiencing social isolation.[54] Such factors may encompass anxiety and depression, kinesiophobia, physical inactivity, and diminished self-efficacy.[55] Given the adverse health consequences of social isolation in individuals with OA and the potential medical benefits for this condition to be reversed, it is crucial to devise and evaluate strategies to effectively address this unmet healthcare need.

### Strengths and limitations of this study

The major strengths of the study include the large sample size from two well-established nationwide cohorts in the UK and US. The findings were generally consistent within the two cohorts, except for the association between moderate alcohol drinking and all-cause mortality. This is the first study to construct an overall healthy lifestyle score to comprehensively evaluate the complex relationships between lifestyle factors and mortality in the OA population. Sensitivity analyses were also conducted to demonstrate the robustness of the findings. Despite these strengths, several limitations are acknowledged. First, the lifestyle information was mainly self-reported, leading to potential measurement errors. Second, the overall healthy lifestyle score was based on the assumption that all lifestyle factors had equal effects on health outcomes, which may not be true. However, the sensitivity analysis using a weighted lifestyle score produced similar results. Third, the follow-up duration was relatively short and those who died during the study period may have had serious diseases at baseline, potentially influencing their lifestyle behaviors. Although the sensitivity analysis adjusting for comorbidities and excluding those with major death-related diseases at baseline generated consistent results, reverse causation and residual confounding cannot be fully eliminated. Fourth, some participants were excluded from the survival analyses due to missing data on lifestyle factors, which may have introduced bias. However, sensitivity analysis using imputed data produced similar results. Fifth, the US NHANES did not have information on social connections. Thus the association between healthy social connections and mortality could not be tested. Sixth, The questionnaire questions and prompts for PA in the two databases are somewhat different, especially UK biobank considers cycling time as part of MPA or VPA, while US NHANES puts it with walking time in the same question, so this may be one of the reasons for the different shapes of the PA RCS in the two databases. However, healhy PA defined by UK biobank was also found to reduce all-cause mortality in US OA population. Finally, the number of participants in some lifestyle categories and events in the US NHANES may be insufficient, and the results should be interpreted with caution.

### Conclusion and public health implications

Based on the UK biobank, the nonlinear relationships suggested patients with OA had the lowest risk of all-cause mortality when BMI was 28 kg/m^2^; sleep was 7 hours/day, VPA was 240 minutes/week, sedentary time was less than 5 hours/day, MPA was more than 550 minutes/week. Additionally, the newly constructed healthy lifestyle score for the OA population combining BMI, diet, sleep duration, physical activity, sedentary time, social connection, smoking and alcohol drinking was associated with a lower risk of all-cause mortality in the UK and US. Our findings first defined the OA-specific healthy lifestyles that can individually and collectively reduce mortality risk.

## Data Availability

The phenotypic UK Biobank data are available on application to the UK Biobank (www.ukbiobank.ac.uk/)

## Contributors

ZZ, FN and DC conceived the study. TF, MZ, QY, YZ, CD, and ZZ contributed to the study design. TF, MZ, and XF processed the data and conducted the main analysis, TF, MZ, XF, YL, PC, ZW, YZ, HC, WH, LL, HG, and DJH wrote the original manuscript draft. TF, XF, SC, and HY undertook simulation and sensitivity analyses. All authors revised the manuscript for important intellectual content, and approved its final version. TF is the guarantor and attests that all listed authors meet authorship criteria and that no others meeting the criteria have been omitted. We thank the UK Biobank participants and the UK Biobank team for generating an important research resource.

## Copyright

The Corresponding Author has the right to grant on behalf of all authors and does grant on behalf of all authors, a worldwide licence to the Publishers and its licensees in perpetuity, in all forms, formats and media (whether known now or created in the future), to i) publish, reproduce, distribute, display and store the Contribution, ii) translate the Contribution into other languages, create adaptations, reprints, include within collections and create summaries, extracts and/or, abstracts of the Contribution, iii) create any other derivative work(s) based on the Contribution, iv) to exploit all subsidiary rights in the Contribution, v) the inclusion of electronic links from the Contribution to third party material where-ever it may be located; and, vi) licence any third party to do any or all of the above.

## Competing interests

All authors have completed the ICMJE uniform disclosure form at http://www.icmje.org/disclosure-of-interest/ and declare: Hunter DJ Provides consulting advice on scientific advisory boards for Pfizer, Lilly, TLCBio, Novartis. QY works in a unit that receives core funding from the University of Bristol and UK Medical Research Council (MM_UU_00011/6).

## Ethical approval

UK Biobank has received ethical approval from the UK national health service’s National Research Ethics Service (16/NW/0274). The present analyses were conducted under UK Biobank application number 67654.

## Funding

The present study was supported by the National Natural Science Foundation of China (32000925), Guangzhou Science and Technology Program (202002030481) and Clinical Research Startup Program of Southern Medical University (LC2019ZD015).

## Notes

### Author Declarations

We used data from the UK Biobank (application number 67654) and US NHANES (2007-2018).

## Reference

1 Hunter DJ, Bierma-Zeinstra S. Osteoarthritis. Lancet 2019; 393:1745–59. doi:10.1016/S0140-6736(19)30417-9

2 Long H, Liu Q, Yin H, et al. Prevalence Trends of Site-Specific Osteoarthritis From 1990 to 2019: Findings From the Global Burden of Disease Study 2019. Arthritis Rheumatol (Hoboken, NJ) 2022;74:1172–83. doi:10.1002/art.42089

3 Palazzo C, Nguyen C, Lefevre-Colau MM, et al. Risk factors and burden of osteoarthritis. Ann Phys Rehabil Med 2016;59:134–8. doi:10.1016/j.rehab.2016.01.006

4 Nüesch E, Dieppe P, Reichenbach S, et al. All cause and disease specific mortality in patients with knee or hip osteoarthritis: Population based cohort study. Bmj 2011;342:638. doi:10.1136/bmj.d1165

5 Wilkie R, Parmar SS, Blagojevic-Bucknall M, et al. Reasons why osteoarthritis predicts mortality: path analysis within a Cox proportional hazards model. RMD Open 2019;5:e001048. doi:10.1136/rmdopen-2019-001048

6 Seale B, Thomas M, Parmar S, et al. 154 Osteoarthritis and premature mortality: pathways for hand, hip, knee and foot osteoarthritis. Rheumatology 2019;58. doi:10.1093/rheumatology/kez108.062

7 van Dam RM, Li T, Spiegelman D, et al. Combined impact of lifestyle factors on mortality: prospective cohort study in US women. BMJ 2008;337:a1440. doi:10.1136/bmj.a1440

8 Hartley L, Dyakova M, Holmes J, et al. Yoga for the primary prevention of cardiovascular disease. Cochrane database Syst Rev 2014;:CD010072. doi:10.1002/14651858.CD010072.pub2

9 Cao Y, Dhana K, Yuan C. Association of a Healthy Lifestyle With All-Cause and Cause-Specific Mortality Among Individuals With Type 2 DiabeteslJ: A Prospective Study in UK Biobank. Published Online First: 2021. doi:10.2337/dc21-1512

10 Veronese N, Li Y, Manson JE, et al. Combined associations of body weight and lifestyle factors with all cause and cause specific mortality in men and women: Prospective cohort study. BMJ 2016;355. doi:10.1136/bmj.i5855

11 Zhang Y-B, Chen C, Pan X-F, et al. Associations of healthy lifestyle and socioeconomic status with mortality and incident cardiovascular disease: two prospective cohort studies. BMJ 2021;373:n604. doi:10.1136/bmj.n604

12 Colpani V, Baena CP, Jaspers L, et al. Lifestyle factors, cardiovascular disease and all-cause mortality in middle-aged and elderly women: a systematic review and meta-analysis. Eur J Epidemiol 2018;33:831–45. doi:10.1007/s10654-018-0374-z

13 Liu Q, Niu J, Huang J, et al. Knee osteoarthritis and all-cause mortality: the Wuchuan Osteoarthritis Study. Osteoarthr Cartil 2015;23:1154–7. doi:10.1016/j.joca.2015.03.021

14 Mendy A, Park J, Vieira ER. Osteoarthritis and risk of mortality in the USA: a population-based cohort study. Int J Epidemiol 2018;47:1821–9. doi:10.1093/ije/dyy187

15 Barbour KE, Lui LY, Nevitt MC, et al. Hip osteoarthritis and the risk of all-cause and disease-specific mortality in older women: A population-based cohort study. Arthritis Rheumatol 2015;67:1798–805. doi:10.1002/art.39113

16 Losina E, Silva GS, Smith KC, et al. Quality-Adjusted Life-Years Lost Due to Physical Inactivity in a US Population With Osteoarthritis. Arthritis Care Res (Hoboken) 2020;72:1349–57. doi:10.1002/acr.24035

17 Sudlow C, Gallacher J, Allen N, et al. UK Biobank: An Open Access Resource for Identifying the Causes of a Wide Range of Complex Diseases of Middle and Old Age. PLoS Med 2015;12:1–10. doi:10.1371/journal.pmed.1001779

18. UK BIOBANK ETHICS AND GOVERNANCE FRAMEWORK Version 3.0 (October 2007). https://www.ukbiobank.ac.uk/media/0xsbmfmw/egf.pdf

19 Wan Z, Guo J, Pan A, et al. Association of Serum 25-Hydroxyvitamin D Concentrations With All-Cause and Cause-Specific Mortality Among Individuals With Diabetes. Diabetes Care 2021;44:350–7. doi:10.2337/dc20-1485

20 Zhang YB o., Chen C, Pan XF, et al. Associations of healthy lifestyle and socioeconomic status with mortality and incident cardiovascular disease: Two prospective cohort studies. BMJ 2021;373. doi:10.1136/bmj.n604

21 Cao C, Friedenreich CM, Yang L. Association of Daily Sitting Time and Leisure-Time Physical Activity With Survival Among US Cancer Survivors. JAMA Oncol 2022;8:395–403. doi:10.1001/jamaoncol.2021.6590

22. Guidelines for Data Processing and Analysis of the International Physical Activity Questionnaire (IPAQ). https://biobank.ndph.ox.ac.uk/ukb/ukb/docs/ipaq_analysis.pdf

23 Cleland C, Ferguson S, Ellis G, et al. Validity of the International Physical Activity Questionnaire (IPAQ) for assessing moderate-to-vigorous physical activity and sedentary behaviour of older adults in the United Kingdom. BMC Med Res Methodol 2018;18:176. doi:10.1186/s12874-018-0642-3

24 Huang SY, Li YZ, Zhang YR, et al. Sleep, physical activity, sedentary behavior, and risk of incident dementia: a prospective cohort study of 431,924 UK Biobank participants. Mol Psychiatry Published Online First: 2022. doi:10.1038/s41380-022-01655-y

25 Chudasama Y V., Khunti K, Gillies CL, et al. Healthy lifestyle and life expectancy in people with multimorbidity in the UK Biobank: A longitudinal cohort study. PLoS Med 2020;17:1–18. doi:10.1371/journal.pmed.1003332

26 Kunzmann AT, Mallon KP, Hunter RF, et al. Physical activity, sedentary behaviour and risk of oesophago-gastric cancer: A prospective cohort study within UK Biobank. United Eur Gastroenterol J 2018;6:1144–54. doi:10.1177/2050640618783558

27 Smith RW, Barnes I, Green J, et al. Social isolation and risk of heart disease and stroke: analysis of two large UK prospective studies. Lancet Public Heal 2021;6:e232–9. doi:10.1016/S2468-2667(20)30291-7

28 Abdullah Said M, Verweij N, Van Der Harst P. Associations of combined genetic and lifestyle risks with incident cardiovascular disease and diabetes in the UK biobank study. JAMA Cardiol 2018;3:693–702. doi:10.1001/jamacardio.2018.1717

29 Liu J, Rehm CD, Onopa J, et al. Trends in Diet Quality Among Youth in the United States, 1999-2016. JAMA 2020;323:1161–74. doi:10.1001/jama.2020.0878

30 Li B, Chen L, Hu X, et al. Association of Serum Uric Acid With All-Cause and Cardiovascular Mortality in Diabetes. Diabetes Care 2023;46:425–33. doi:10.2337/dc22-1339

31 Lee DH, Keum N, Hu FB, et al. Predicted lean body mass, fat mass, and all cause and cause specific mortality in men: prospective US cohort study. BMJ 2018;362:k2575. doi:10.1136/bmj.k2575

32. World Health Organization. Ageing and health. 2018.

33 Lourida I, Hannon E, Littlejohns TJ, et al. Association of Lifestyle and Genetic Risk With Incidence of Dementia. JAMA 2019;:1–8. doi:10.1001/jama.2019.9879

34 Van Buuren S, Groothuis-Oudshoorn K. mice: Multivariate imputation by chained equations in R. J Stat Softw 2011;45:1–67.

35 Sliepen M, Mauricio E, Lipperts M, et al. Objective assessment of physical activity and sedentary behaviour in knee osteoarthritis patients – beyond daily steps and total sedentary time. BMC Musculoskelet Disord 2018;19:64. doi:10.1186/s12891-018-1980-3

36 Rausch Osthoff AK, Niedermann K, Braun J, et al. 2018 EULAR recommendations for physical activity in people with inflammatory arthritis and osteoarthritis. Ann Rheum Dis 2018;**77**:1251–60. doi:10.1136/annrheumdis-2018-213585

37. Physical Activity Guidelines for Americans, 2nd edition.

38 Yang YJ. An Overview of Current Physical Activity Recommendations in Primary Care. Korean J Fam Med 2019;40:135–42. doi:10.4082/kjfm.19.0038

39 Corrigendum to: 2021 ESC Guidelines on cardiovascular disease prevention in clinical practice: Developed by the Task Force for cardiovascular disease prevention in clinical practice with representatives of the European Society of Cardiology and 12 medical. Eur Heart J 2022;**43**:4468. doi:10.1093/eurheartj/ehac458

40 Salis Z, Gallego B, Nguyen T V, et al. Decrease in body mass index is associated with reduced incidence and progression of the structural defects of knee osteoarthritis: a prospective multi-cohort study. Arthritis Rheumatol (Hoboken, NJ) Published Online First: 16 August 2022. doi:10.1002/art.42307

41 Reijman M, Pols HAP, Bergink AP, et al. Body mass index associated with onset and progression of osteoarthritis of the knee but not of the hip: the Rotterdam Study. Ann Rheum Dis 2007;66:158–62. doi:10.1136/ard.2006.053538

42 Flegal KM, Kit BK, Orpana H, et al. Association of all-cause mortality with overweight and obesity using standard body mass index categories: a systematic review and meta-analysis. JAMA 2013;309:71–82. doi:10.1001/jama.2012.113905

43 Prospective Studies Collaboration, Whitlock G, Lewington S, et al. Body-mass index and cause-specific mortality in 900 000 adults: collaborative analyses of 57 prospective studies. Lancet (London, England) 2009;373:1083–96. doi:10.1016/S0140-6736(09)60318-4

44 Mozaffarian D. Dietary and Policy Priorities for Cardiovascular Disease, Diabetes, and Obesity: A Comprehensive Review. Circulation 2016;133:187–225. doi:10.1161/CIRCULATIONAHA.115.018585

45 Said MA, Verweij N, van der Harst P. Associations of Combined Genetic and Lifestyle Risks With Incident Cardiovascular Disease and Diabetes in the UK Biobank Study. JAMA Cardiol 2018;3:693–702. doi:10.1001/jamacardio.2018.1717

46 Svensson T, Saito E, Svensson AK, et al. Association of Sleep Duration With All- and Major-Cause Mortality Among Adults in Japan, China, Singapore, and Korea. JAMA Netw open 2021;4:e2122837. doi:10.1001/jamanetworkopen.2021.22837

47 da Silva AA, de Mello RGB, Schaan CW, et al. Sleep duration and mortality in the elderly: a systematic review with meta-analysis. BMJ Open 2016;6:e008119. doi:10.1136/bmjopen-2015-008119

48 Mnatzaganian G, Ryan P, Norman PE, et al. Smoking, body weight, physical exercise, and risk of lower limb total joint replacement in a population-based cohort of men. Arthritis Rheum 2011;63:2523–30. doi:10.1002/art.30400

49 Leung YY, Ang LW, Thumboo J, et al. Cigarette smoking and risk of total knee replacement for severe osteoarthritis among Chinese in Singapore--the Singapore Chinese health study. Osteoarthr Cartil 2014;22:764–70. doi:10.1016/j.joca.2014.03.013

50 Gullahorn L, Lippiello L, Karpman R. Smoking and osteoarthritis: differential effect of nicotine on human chondrocyte glycosaminoglycan and collagen synthesis. Osteoarthr Cartil 2005;13:942–3. doi:10.1016/j.joca.2005.03.001

51 Dashti HS, Miranda N, Cade BE, et al. Interaction of obesity polygenic score with lifestyle risk factors in an electronic health record biobank. BMC Med 2022;20:5. doi:10.1186/s12916-021-02198-9

52 Tanskanen J, Anttila T. A Prospective Study of Social Isolation, Loneliness, and Mortality in Finland. Am J Public Health 2016;106:2042–8. doi:10.2105/AJPH.2016.303431

53 Alcaraz KI, Eddens KS, Blase JL, et al. Social Isolation and Mortality in US Black and White Men and Women. Am J Epidemiol 2019;188:102–9. doi:10.1093/aje/kwy231

54 Siviero P, Veronese N, Smith T, et al. Association Between Osteoarthritis and Social Isolation: Data From the EPOSA Study. J Am Geriatr Soc 2020;68:87–95. doi:10.1111/jgs.16159

55 Ethgen O, Vanparijs P, Delhalle S, et al. Social support and health-related quality of life in hip and knee osteoarthritis. Qual Life Res 2004;13:321–30. doi:10.1023/B:QURE.0000018492.40262.d1

